# Wastewater Metagenomic Virome Analysis of the 2026 World Cup in Texas

**DOI:** 10.64898/2026.09.04.26362178

**Authors:** Ryan King Perez, Harihara Prakash, Dustin Jaynes, Phil Huang, Jennifer Deegan, Loren Hopkins, Matt Ross, Marissa Minor, Katelyn Payne, Tulin Ayvaz, Justin R. Clark, Eric Boerwinkle, Michael J. Tisza, Anthony W. Maresso

## Abstract

Mass gatherings are thought to create conditions for pathogen introduction and spread, yet whether such events measurably alter the viral community of a city has rarely been tested against an extensive historical baseline. We analyzed 5,137 hybrid-capture metagenomic wastewater samples collected from 55 sites across 17 Texas cities between June 2022 and July 2026, spanning the 2026 FIFA World Cup. We asked (1) whether host-city viral community composition during the tournament departed from its established summer-to-summer range, (2) whether targeted respiratory viruses rose above their characteristic summer trend, and (3) whether the load of rare and novel taxa was elevated in host cities relative to non-host cities. Despite the plausibility of these hypotheses, we detected no tournament-associated perturbations. Although based on a single event, these findings favor continuous wastewater surveillance over event-triggered deployment, both for detecting introductions that arrive unpredictably and for supplying the baseline against which any event should be judged.

## Introduction

The 2026 FIFA World Cup was a global event that brought an estimated 3 million visitors and travelers across North American host cities, including Houston and Dallas (Angelo, 2025). The tournament had a fixed window (June 11^th^ through July 19^th^, 2026) occurring in specific host cities, which allowed a rare natural experiment: a large, time-boxed influx of people into defined locations, against a backdrop that otherwise stayed the same. Since it is commonly thought that the sudden influx of travelers to an otherwise stable location may also bring a concomitant increase in pathogens, we tested this hypothesis against a long-running viral monitoring program in the cities experiencing these events.

Despite the recognized risk of mass gatherings, few studies have been able to ask whether a large global event shifts the viral community of a city (Memish et al, 2019). Most surveillance is clinical, event-specific, lacks a long-term baseline to judge what counts as unusual, or uses targeted approaches that only assess one or a few viral pathogens. Over the past 4.5 years, our team has been running a weekly wastewater surveillance program across many Texas cities and sites, generating greater than 5,000 sequenced samples in the process (Clark et al, 2023). This program, TexWEB (Texas Wastewater and Environmental Biomonitoring), unlike other wastewater monitoring programs, uses wastewater metagenomics, *i*.*e*. semi-agnostic sequencing of up to 3,000 different human viruses from a given sample. The program has had highly impactful and headlined viral detections, including characterizing hundreds of viruses not known to be in wastewater, avian flu, measles, HIV, oncoviruses, parvovirus B19 outbreaks and more (Tisza et al, 2023; Tisza et al, 2024; Javornik Cregeen et al, 2025; Clark et al, 2026; Harihara et al, 2026; Clark et al, 2025; Bauer et al, 2026).

TexWEB’s ongoing analysis and the fortuitous placement of World Cup events in cities enrolled in the program afforded a unique opportunity to conduct an analysis of the wastewater virome using data before, during, and after the World Cup in Texas host cities (Dallas and Houston). In addition, because we have years of historic data at these sites and data from nearby metros not hosting events, we can assess if there are perturbations in the viral community in a more controlled manner. We hypothesized that an influx of international travelers, including large groups arriving from regions in their own respiratory-virus season, would introduce uncharacteristic pathogens and shift the community composition of host-city wastewater beyond its normal year-to-year or summer-to-summer range. Unexpectedly, our data suggest that large-scale shifts in the virome did not occur. Reasons for this are offered but it may suggest that the maximum benefit afforded by wastewater analysis are best achieved through continuous monitoring instead of focused event driven activities.

## Results

### The virome remained within interannual variation during the 2026 World Cup

Between June 2022 and July 2026, the TexWEB program collected over 5,000 samples across 17 Texas cities, from 55 sites. Sampling effort was distributed with the greatest number of samples collected in Houston and El Paso, alongside shallower collection counts at more recently enrolled cities with fewer sites (Figure 1A, 1B). Across this collection, hybrid-capture sequencing recovered viral taxa spanning more than 20 orders and both DNA and RNA virus phyla (Figure 1C), with node color reflecting each clade’s abundance restricted to samples collected during the World Cup tournament window (June 11^th^ to July 19^th^, 2026). The color gradient highlights tournament-window viral activity against the full Texas viral landscape, with node size showcasing the prevalence of the given phyla for the entire multi-year collection. Weekly sampling of wastewater sites continued through the tournament window across all cities, providing contemporaneous coverage of all Texas match days (Figure 1D). As perhaps the longest continuous metagenomic wastewater surveillance program known to the authors, this multi-year sampling allowed us to characterize the range of summer-to-summer viral community variation at several sites prior to the 2026 FIFA World Cup.

**Figure 1.**
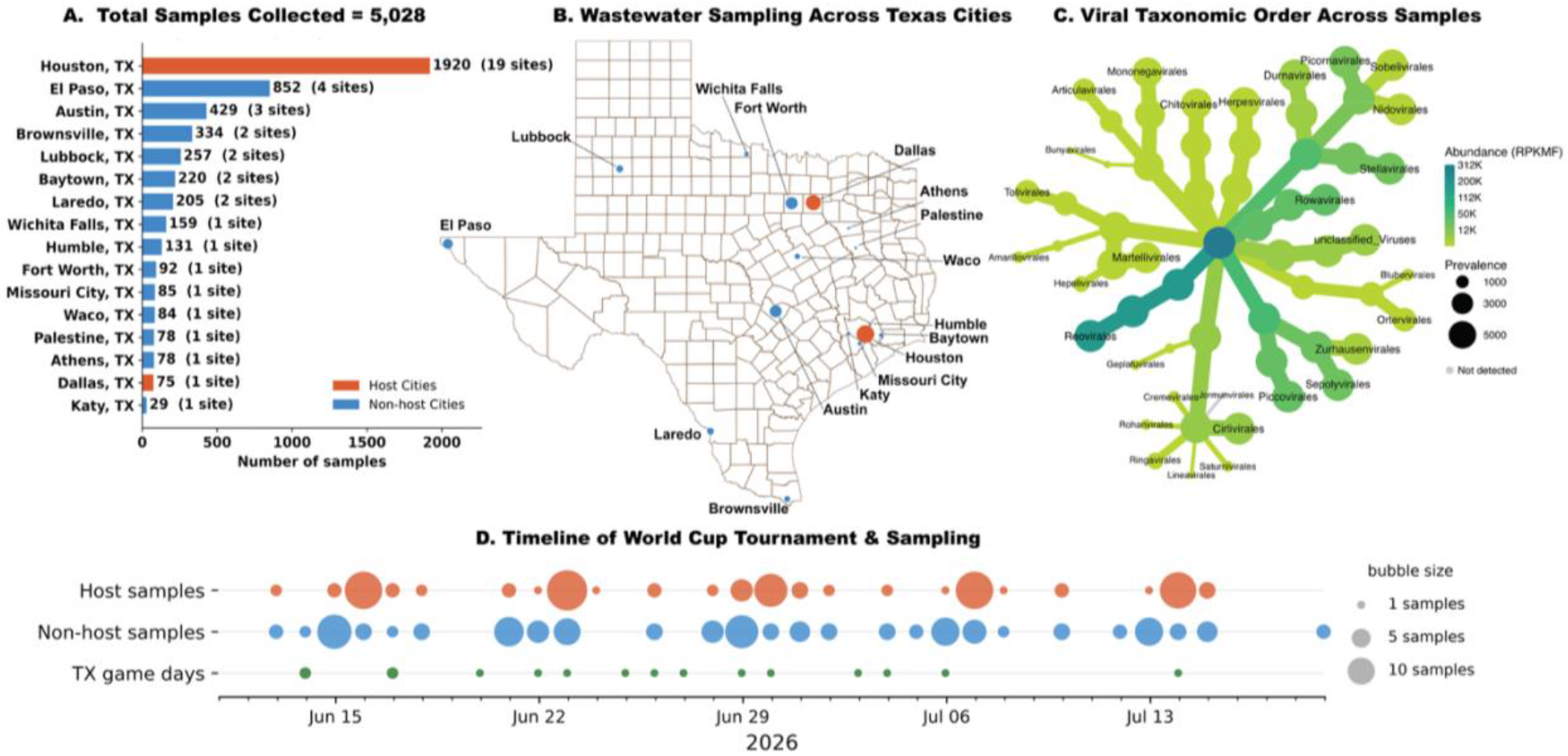
Overview of wastewater samples across Texas cities from May 2022 through July 2026. **(A)** The total wastewater samples collected from each city, with the number of sites per city in parentheses. Note: for Dallas, we merged Arlington samples with Dallas to be consistent with the FIFA World Cup naming conventions. **(B)** A geographical layout of the population served in 2025 (most recent Census information) within the TexWEB wastewater sequencing program. Each bubble is proportional to the total population for each city. **(C)** A heat tree showing viral abundance of samples during the World Cup tournament (gradient coloring, RPKMF) and prevalence of viruses across all samples collected since the start of the TexWEB wastewater program (thickness of the branch). **(D)** A timeline visualizing the distribution across time for: (top line) the number of host-city samples collected, (middle line) non-host-city samples collected, and (bottom line) the number of World Cup matches played on a given day.

Since this baseline requires several years of continuous same-site collection, Houston was the sole host city with sufficient historical depth to support this comparison. Therefore, we focus this section of the analysis on Houston’s six long-running sites. This set the baseline against which any tournament-associated perturbation could be compared. Starting with sites with four or more years of collected samples, we compared viral community composition across June-July intervals in Houston (2022 through 2026). We first asked whether summers were distinguishable from one another at a given site. Communities varied significantly by summer (Figure 2A; PERMANOVA p=0.001 at all six sites), confirming the sensitivity of our approach successfully resolving year-to-year compositional shifts. We hypothesized that the influx of visitors would shift host-city composition during the World Cup tournament window beyond this interannual range. To test this, we quantified how compositionally distinct Summer 2026, the World Cup period, was compared to historical summers, relative to how distinct historical summers are from each other. For each site analyzed, we computed the Bray-Curtis dissimilarity between cross-summer sample pairs and summarized each summer pair by its median distance. Then, we compared Summer 2026 versus each historical summer and measured if that had a larger compositional shift than the given sites historical summer-vs-summer paired comparisons. If we saw a perturbation from the tournament beyond normal interannual variations, the Summer 2026 comparisons should sit significantly higher than the historical baseline comparisons. Pairwise Bray-Curtis comparisons of Summer 2026 to each historical summer were indistinguishable from the historical summer-to-summer pairwise baseline (Mann-Whitney U-test, one-sided, p > .05 at all 6 sites, Holm-corrected; Figure 2B, Supplemental Table 1). Additionally, effect sizes varied across sites (rank-biserial r = 0.08 to 0.92), with J6, S3, and Z9 sites showing the largest separation (Supplemental Table 1). Importantly, two of our sites were from catchment areas containing an international airport. Comparison of the World Cup window to past summers of the same time frame for these airports also did not yield noticeable differences in virome diversity (data not shown). Taken together, this data suggest the tournament produced no detectable compositional shift beyond normal interannual variation.

**Figure 2.**
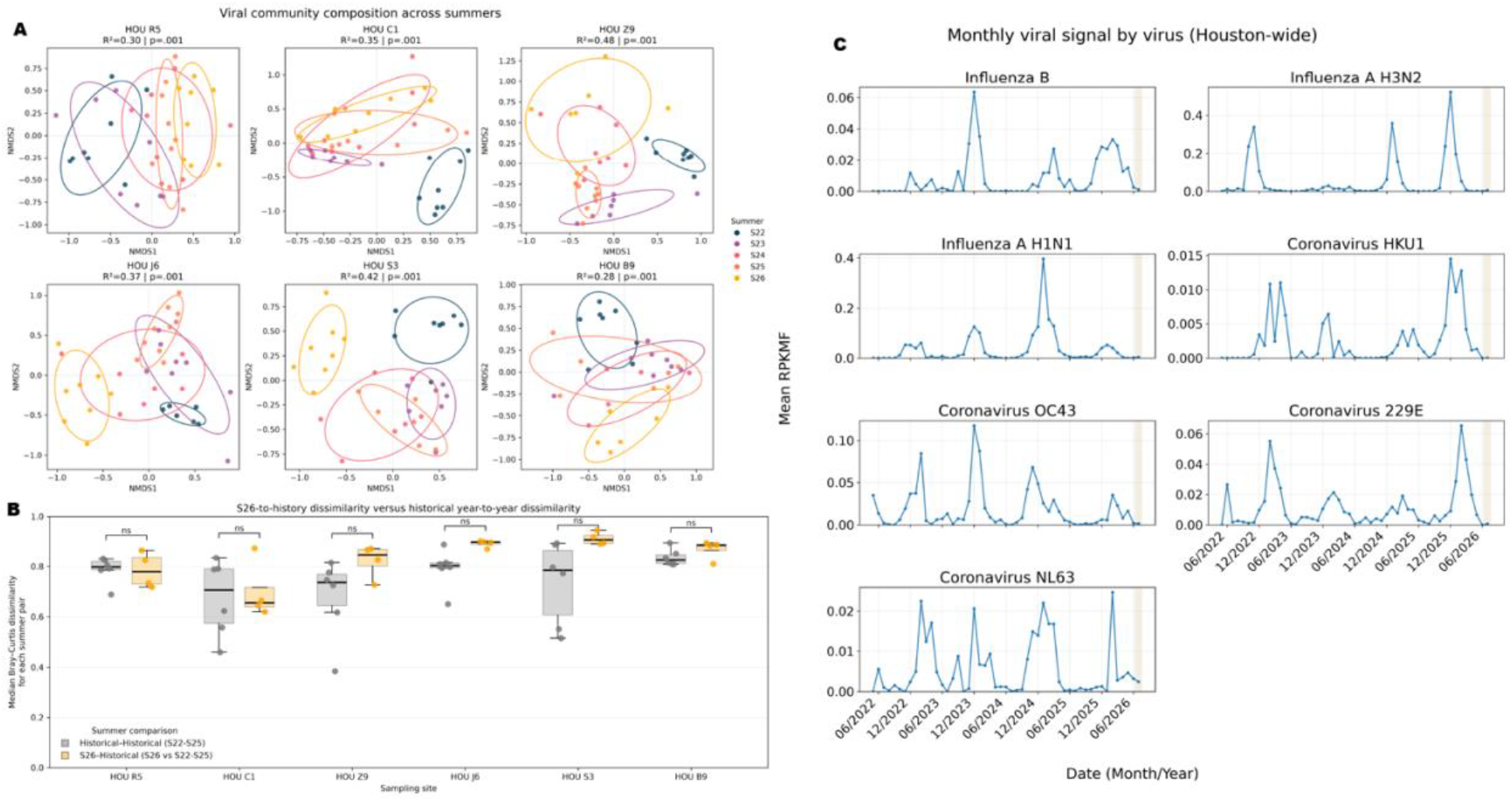
Summer analysis of Houston viral community composition, 2022-2026. **(A)** Non-metric multidimensional scaling (NMDS) ordinations of Bray-Curtis dissimilarity at each of the six Houston sites (HOU R5, C1, Z9, J6, S3, B9), with samples colored by summer (S22-S26) and 95% ellipses per summer. Communities were significantly distinguishable by summer at every site (PERMANOVA, 999 permutations; R^2^ annotated per panel, R^2^ ranging from 0.28-0.48; p=0.001 for all six sites). **(B)** Per site, the median Bray-Curtis dissimilarity of cross-summer sample pairs, split into historical summer-vs-summer comparisons (S22-S25; gray) and Summer 2026-vs-historical comparisons (yellow). Summer 2026 was indistinguishable from the historical baseline at all six sites (one-sided Mann-Whitney U, Holm-corrected; n.s.; Supplemental Table 1), indicating no detectable compositional shift beyond interannual variation. **(C)** Monthly mean relative abundance (RPKMF), Houston-wide, of targeted respiratory viruses. The shaded band marks the tournament window. n.s. denotes not significant.

### Off-season respiratory viruses showed no rise despite international visitor influx

Respiratory viruses such as influenza and the seasonal human coronaviruses follow a predictable seasonal pattern of increased abundance during each winter and fall to low troughs over each summer (Kakoullis et al, 2023). Our sequencing approach reliably quantifies these viruses across the full seasonal range, including the low-abundance shoulder months flanking each winter peak. This demonstrates sensitivity to small, real respiratory-virus signals well below peak levels. Since the Southern Hemisphere experiences its own respiratory-virus season during the Northern Hemisphere’s summer, the large influx of international visitors to Houston during the June through July tournament could have plausibly imported these viruses during the off-season (Kakoullis et al, 2023). This would elevate their abundance above the low summer baseline we normally observe. Instead, across five years of summers, each virus (namely influenza B, influenza A H3N2, influenza A H1N1, coronavirus HKU1, coronavirus OC43, coronavirus 229E, and coronavirus NL63) tracked its expected seasonal cycle, with pronounced winter peaks collapsing to low summer troughs (Figure 2C). During the 2026 World Cup window, abundance remained at the characteristic summer lows at every Houston site with no off-season rise detectable (Supplemental Figure 1).

### Rare and novel viruses were not enriched in World Cup Host Cities

We used the broad nature of our methodology (enriching for over 3,000 viral species) to then ask whether the tournament altered the load of unusual taxa. We looked at all samples collected across Texas cities, where at least 26 samples were collected (equating to six months’ worth of sampling or longer) to set an established baseline. From samples collected pre-tournament, we defined two-groups: rare taxa (defined as a virus detected in less than or equal to 10 pre-tournament samples, picked from a detection range of thresholds, Supplemental Figure 2) and novel taxa (viruses absent from the pre-tournament sampling entirely but appearing during the tournament window). Then, we scored every sample by its combined count of rare and novel taxa (termed “unique” taxa), comparing host and non-host cities over time (Figure 3).

**Figure 3.**
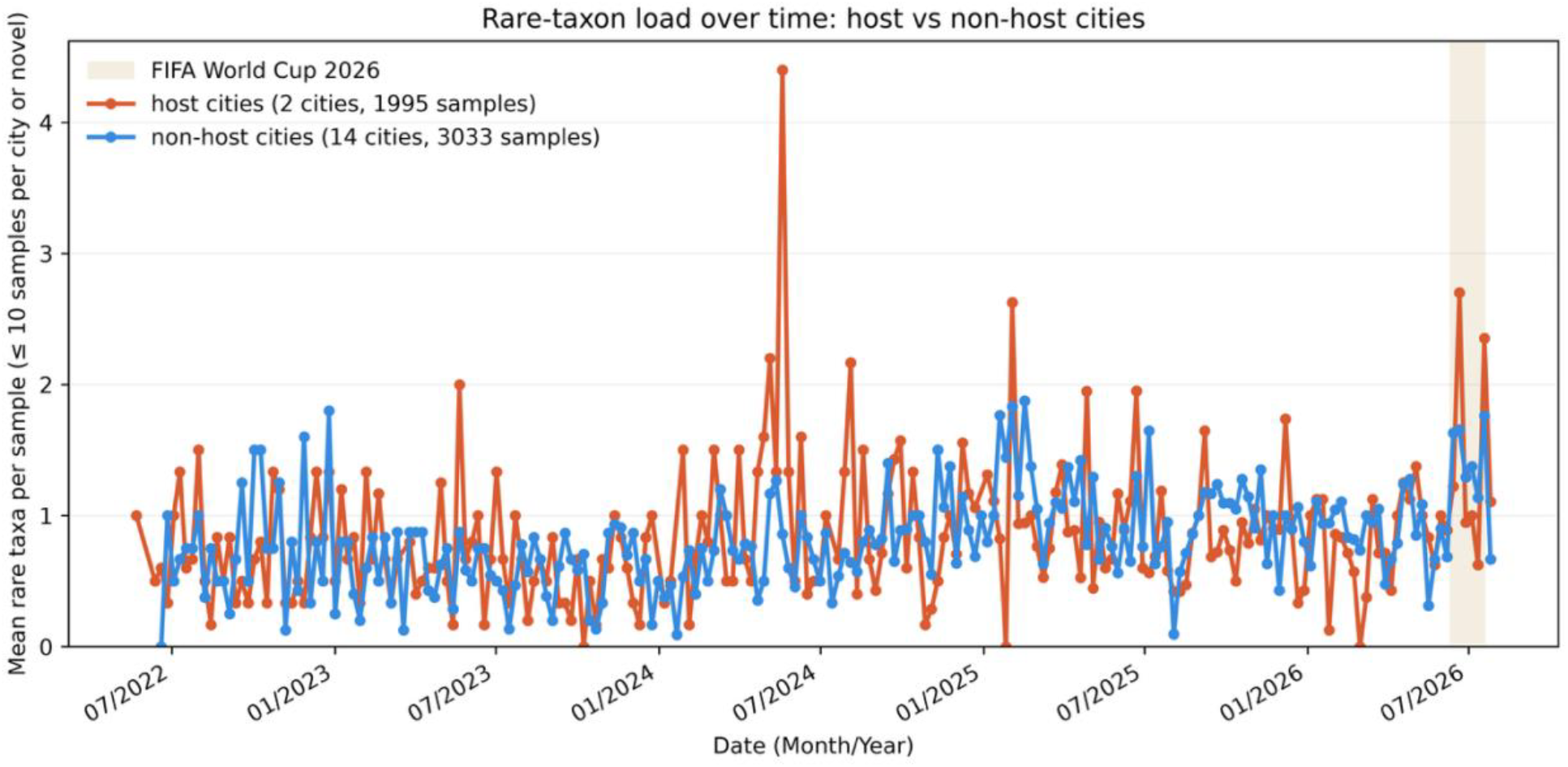
Rare-taxon load over time in host versus non-host Texas cities. Weekly mean count of “unusual” taxa per sample: rare taxa (viruses detected in less than or equal to 10 baseline/pre-tournament samples) plus novel taxa (viruses absent from all baseline samples but appearing afterward) for host cities (Houston and Dallas; 2 cities, 1,995 samples; coral) and non-host Texas cities (14 cities, 3,033 samples; blue). The shaded band marks the 2026 FIFA World Cup window. Host cities showed no elevation relative to non-host cities during the window (two-sided Mann-Whitney U, p=.76), and the visual spikes at the window’s leading edge fell within normal between-city variation.

During the tournament window, host cities showed no elevation in unique taxa load relative to non-host cities across Texas (Mann-Whitney U, p >= .05, two-sided, Supplemental Table 2). The apparent visual spikes at the tournament window’s leading edge (Figure 3) fell within the normal range of variation between host and non-host cities. We then asked whether host cities were elevated relative to their own historical summers, comparing the 2026 window samples to the same calendar span in 2022 through 2025. We did find a significant difference in rare-taxa load in 2026 exceeding prior summers (p < .01, Mann-Whitney U two-sided, Supplemental Table 3). However, this elevation was not specific to host cities as non-host cities rose in parallel. The 2026 increase suggests a state-wide shift rather than a tournament effect concentrated where visitors gathered. The source of this broad rise may reflect a genuine ecological shift in the regional virome.

## Discussion

Mass gatherings elevate the conditions under which a pathogen can be introduced in a community and circulate before clinical systems register it. Yet whether such an event measurably alters the viral makeup of a host city has rarely been tested, in part because doing so requires a longitudinal baseline against which unusual can be defined (Kitajima et al, 2022). The 2026 FIFA World Cup offered that opportunity: an estimated three million visitors converging on North American host cities within a fixed window, against a background that otherwise remained relatively unchanged. Houston hosted seven matches, with a projected influx of roughly 500,000 visitors (Kendrick, 2026). Four years of continuous pre-tournament sequencing at six long-running Houston sites defined the city’s normal summer-to-summer range. We therefore asked whether the tournament shifted host-city wastewater viral community composition, or elevated pathogen load, beyond the range defined by that baseline.

Viral communities at the six long-tested Houston sites were significantly distinguishable by summer (PERMANOVA, p=0.001), confirming our method resolves genuine year-to-year shifts. Against that resolution, Summer 2026 was statistically indistinguishable from the historical summer-to-summer baseline at all six sites tested (Mann-Whitney U, n.s., Holm-corrected). For respiratory Winter-season pathogens, our sequencing approach reliably quantifies influenza and the seasonal human coronaviruses across the full seasonal spectrum, including the low-abundances flanking each Winter peak. Despite an influx of visitors from regions such as South America in their own respiratory-virus season, each targeted virus tracked its expected cycle and remained at characteristic summer lows throughout the tournament window at each Houston site. Broadening beyond targeted taxa, host cities showed no elevation in unique taxon load relative to non-host Texas cities during the tournament window (Mann-Whitney U, p=0.76). The one significant signal we detected was temporal rather than spatial: rare-taxon load in 2026 exceeded the same calendar span in 2022-2025 (Mann-Whitney U, p=0.0095), though non-host cities rose concomitantly, identifying this as a state-wide shift rather than a tournament effect.

Interpreting the null results requires understanding the magnitude of signal our approach can detect. There are thus several limitations in our study. Firstly, Bray-Curtis dissimilarity uses relative abundance, so change in visitor numbers alone might not move community composition. A detectable shift requires that travelers carry viruses distinct from those already circulating and at sufficient prevalence to be detected against the virome signal from city residents. Secondly, the wastewater signal is diluted across the population served by each treatment plant rather than the city. As visitors were not distributed evenly across these catchment sites, with attendance concentrating near the stadium, downtown lodging, and fan festival venues, some of our six sites may not have had the total number of visitors expected. We also acknowledge that we do not know the contribution of population size from the catchment and how that affects the results here in. Thirdly, the tournament window may not have aligned with when an introduced infection would become detectable: a traveler infected in their home country could introduce detectable signals of the newly presented virus days after the sampling window would capture the signal. Despite these potential constraints, the sensitivity of the platform has been historically well-tested detecting measles from two wastewater treatment sites in Texas with reported cases of measles in two unvaccinated adults (Javornik Cregeen et al, 2025) among tens of thousands living in the catchment area, and environmental surveillance has repeatedly identified poliovirus circulating in the absence of clinical cases (Manor et al, 1999; Lesenfants et al, 2026). This shows that low-prevalence introductions are detectable in wastewater when they do occur. Our nulls therefore likely reflect the absence of a community-scale signal rather than the complete absence of any introduction.

During the tournament window, host cities did not have a greater unique taxa load than non-host cities. Our definition of “novel” reflects absence from the pre-tournament sampling baseline and what is present in the reference database, a unique feature our program brings due to the historical data collected. Several explanations could produce a state-wide rise, including sequencing depth drift across the collection samples, changes in the sites and cities contributing samples over time, or a genuine ecological shift in the regional virome. 33 taxa were absent from all pre-tournament samples and detected during the tournament with 30 of these appearing in a single sampling. The list is dominated by agricultural, environmental, and animal-associated viruses, including six genomoviruses and several plant viruses, while only three are human-associated (human erythrovirus V9, Alphapolyomavirus terdecihominis, and Enterovirus B77), each detected once (Supplemental Table 4). Had international travelers seeded novel human pathogens into the Houston wastewater, we could expect this set to be enriched for human viruses recurring across samples and sites throughout the tournament. Instead, the data shows the ordinary rare diversity tail of the virome and may represent normal seasonal diversity entry during summer periods.

These results highlight the complementary role of community-composition and presence-based surveillance. Bray-Curtis dissimilarity is sensitive to changes in abundant taxa, whereas presence analysis displays low-abundance viruses that may not measurably shift overall community composition. The two approaches answer different questions and provide orthogonal observations for the perturbation of the wastewater virome, thus strengthening our conclusions. Studies designed to detect pathogen importation at mass gatherings should consider both presence-based and community-composition metrics. Broad approaches such as large-scale hybrid-capture sequencing provide an important advantage for mass-gathering surveillance by enabling detection of unexpected viral signals beyond a predefined pathogen list. Virus-specific assays like PCR, while powerful, can lose sensitivity to lineages of greatest concern if those lineages diverge from the reference in the primer-binding regions. For example, when Omicron emerged, point mutations under the primers of the standard SARS-CoV-2 amplicon scheme (ARTIC) knocked out the very segments carrying the lineage-defining mutations, recovered only after the primers were redesigned (Ulhuq et al, 2023). On the other hand, hybrid capture is robust to small numbers of mutations since the longer oligos tolerate mismatch during annealing (Kuchinski et al, 2022). Sequencing-based approaches also are comprehensive, capturing potentially thousands of viruses in a single sample, thereby allowing for community composition to be measured at a large-scale.

The absence of a tournament signal or perturbation argues for continuous rather than event-triggered surveillance. The 2026 Bundibugyo outbreak in the Democratic Republic of the Congo illustrates this example. Specimens from the initial cases tested negative on a Zaire-ebolavirus specific PCR assay, the platform in routine regional use (Sullivan, 2026). Bundibugyo virus was not confirmed until three weeks after the index case developed symptoms when researchers ultimately used hybrid capture-based sequencing (Sullivan, 2026). Surveillance organized solely around scheduled events would have been active in Houston during the World Cup and likely found nothing significant, while missing the introductions that really matter such as recent measles or H5N1 outbreaks that did not coincide with mass gatherings but instead were detected as a part of our routine reporting (Tisza et al, 2024). The value of continuous wastewater metagenomics lies in maintaining the multi-year surveillance that makes an anomaly recognizable when one appears.

## Methods

### Sample Collection, Processing, and Sequencing

Wastewater samples were collected, shipped, processed, and sequenced as described by Tisza et al (2023) and Prakash et al (2026). 24-hour composite grab samples were shipped on ice immediately after acquisition and arrived within one day of shipment. Samples were processed within several hours of receipt as outlined in Tisza et al 2023. Raw wastewater was collected across TexWEB sites in Texas, processed at the Alkek Center for Metagenomics and Microbiome research (CMMR, at Baylor College of Medicine), and subjected to nucleic acid extraction, reverse transcription, and Twist hybrid-capture library preparation (with Twist Comprehensive Viral Research Panels) as discussed in Tisza et al (2023). Libraries were sequenced on the Illumina NextSeq platform to generate 150-bp paired-end reads. Reads were quality-filtered with fastp v0.23.2 (Chen et al, 2018) and classified against EsViritu’s Virus Pathogen Database (EsViritu v1.1.6) (Tisza et al. 2023), yielding per-sample relative abundance in RPKMF (Reads Per Kilobase of reference genome per Million reads passing Filtering). RPKMF is a semi-quantitatiive relative abundance measurement of viral reads that scale according to viral concentrations (Tisza et al, 2023; Tisza et al, 2024; Prakash et al, 2026).

### Sample-level quality control and inclusion criteria

We restricted community-composition analysis to sites with samples more than 26 weeks prior to the tournament (corresponding to six months of weekly collection) to ensure a stable longitudinal baseline. Of the 20 cities in the collection, 16 met these criteria and were carried forward in this analysis. We note that Arlington was merged with the city of Dallas, to stay consistent with the FIFA World Cup tournament’s naming conventions. Baseline was defined as all qualifying samples collected before the tournament start (June 11th, 2026), while the tournament window was defined as June 11th to July 19th, 2026.

### Community composition analysis

Per-sample taxonomic profiles were summarized at the strain level. Abundances were row-normalized to relative proportions per sample, and pairwise Bray-Curtis dissimilarities were computed between all samples (scipy v1.18.0). For each of our six long-running Houston sites, summers were defined as June 1 through July 31 of each year (S22-S26). To test whether summers were compositionally distinguishable, we ran PERMANOVA (scikit-bio v0.7.3; 999 permutations) on the Bray-Curtis matrix with summer as the grouping factor, and visualized structure with non-metric multidimensional scaling (NMDS; scikit-learn MDS, metric=False). To test whether Summer 2026 was an outlier, we computed the median Bray-Curtis dissimilarity of all cross-summer sample pairs for each summer pair per site, then compared the distribution of S26-vs-historical medians against the distribution of historical-vs-historical medians using a one-sided Mann-Whitney U test (S26 > historical), with Holm correction across the six sites (statsmodels v0.14.6).

### Seasonal respiratory virus analysis

We evaluated targeted respiratory pathogens by tracking strain-level relative abundance (RPKMF) of influenza (H3N2, H1N1, influenza B) and the seasonal human coronaviruses (HKU1, OC43, 229E, NL63) across Houston samples. Per-sample abundances were aggregated into monthly means and plotted as per-virus time series spanning the full collection period, with the tournament window annotated.

### Rare and novel taxon analysis

To assess unusual-taxon load beyond targeted viruses, we defined two taxon sets from baseline (pre-tournament) samples. Rare taxa were those detected in ≤10 baseline samples; novel taxa were those absent from all baseline samples but appearing afterward. Each sample was scored by its combined count of rare and novel taxa (“unique” taxa). We compared this score between host and non-host cities during the tournament window (two-sided Mann-Whitney U, host > non-host) and separately compared host-city window samples against host-city samples from the same calendar span (June 11-July 19) of 2022-2025 pooled (two-sided Mann-Whitney U, 2026 > historical). The temporal comparison used rare-taxon counts only, since novel taxa are structurally absent from baseline and would inflate the 2026 group.

### Data analysis and visualization of wastewater viruses

All analyses were performed in Python v3.13.14 using pandas v3.0.3, numpy v2.5.0, scipy v1.18.0, scikit-bio v0.7.3, scikit-learn v1.9.0, and statsmodels v0.14.6. Figures were generated with matplotlib v3.11.0 and seaborn v0.13.2.

## Data Availability

All data produced in the present study are available upon reasonable request to the authors.

## Acknowledgements

This work was supported by S.B. 1780, 87th Legislature, 2021 Reg. Sess. (Texas 2021) under the TEPHI program, NIH/NIAID (grant number U19 AI157981), Baylor College of Medicine Joseph Melnick Seed Funds, and Alkek Foundation Seed Funds. The funders of the study had no role in study design, data collection, data analysis, data interpretation, or writing of the research article. We wish to thank the Texas Department of State Health Services for the coordination of samples throughout the World Cup activities, including Heidi Bojes. We would like to thank Dr. Bryan Brooks (Baylor University) and his team for their leadership in wastewater surveillance during the World Cup activities in Texas. This project was also supported by the Centers for Disease Control and Prevention of the U.S. Department of Health and Human Services (HHS) as part of a financial assistance award. The contents are those of the author(s) and do not necessarily represent the official views of, nor an endorsement, by CDC/HHS, or the U.S. Government. We are grateful for the support of the local water utilities and public health professionals, without whom this work would not be possible.

## Supplemental Material

**Figure S1.**
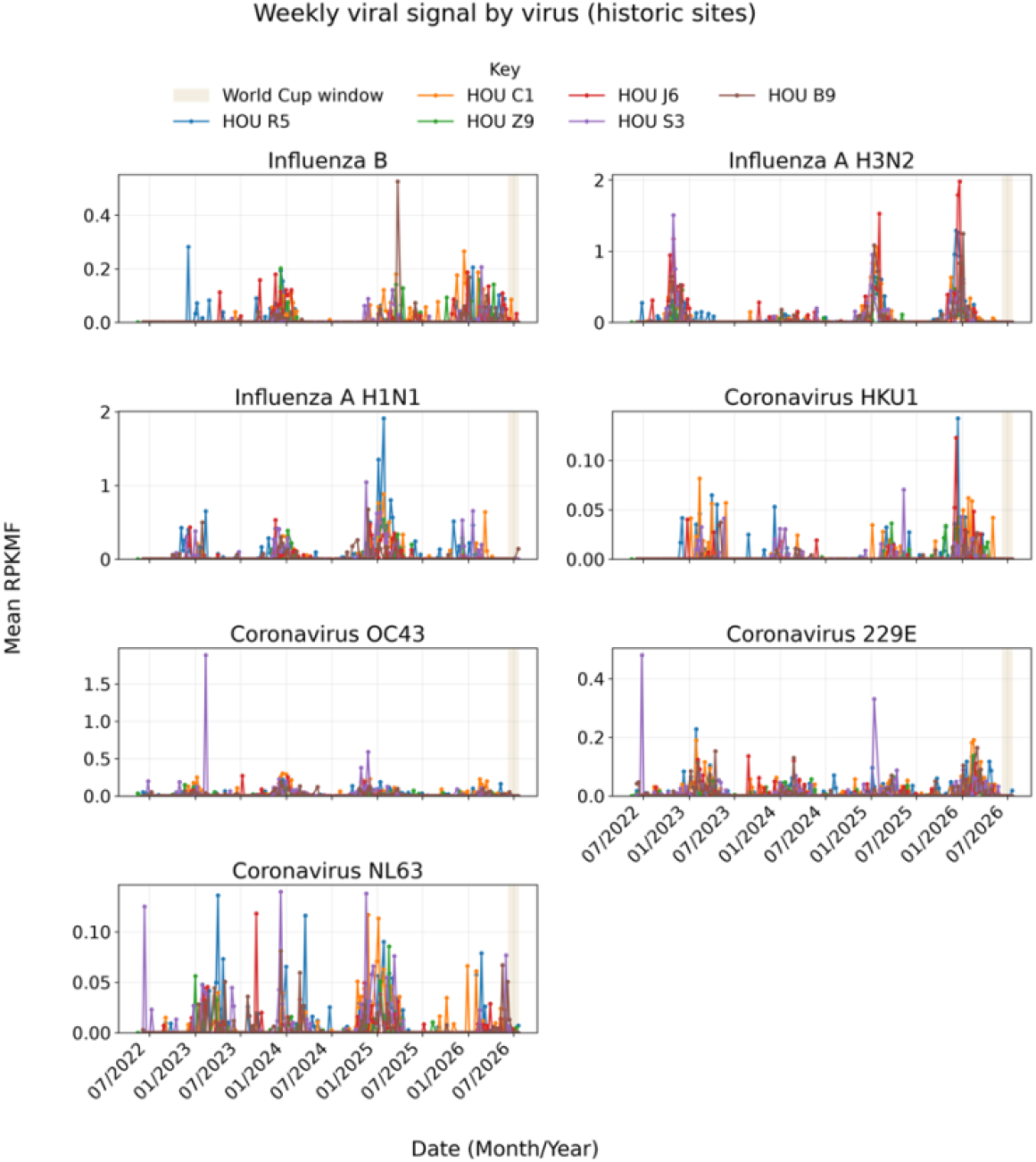
Weekly viral abundance of respiratory viruses at each of the six historic sites in Houston spanning from 2022 to 2026. Time-series plots of the seven seasonal viral abundances (RPKMF) per week since the start of the TexWEB program. Each color represents a specific Houston site, and the shaded band marks the tournament window.

**Figure S2.**
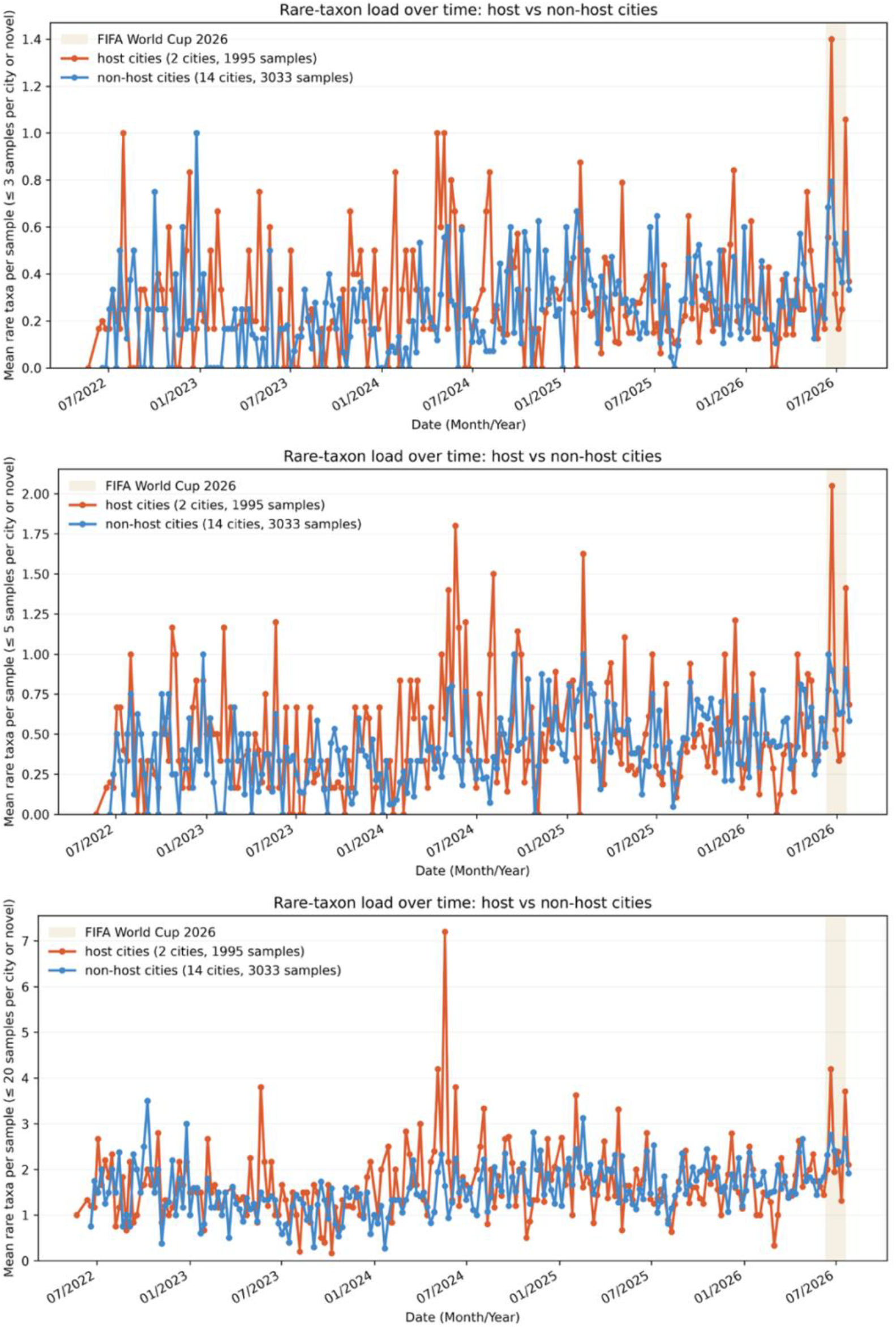
Time-series plot of rare-taxon load between host and non-host cities at thresholds of 3, 5, and 20 viruses. Weekly mean count of unusual taxa per sample, at thresholds of 3 viruses for the top panel, 5 viruses at the center panel, and 20 viruses for the bottom panel. Unusual taxa represents both rare and novel taxa during the world cup tournament window. The shaded band marks the 2026 FIFA World Cup window.

Supplemental Table 1: Per-site Mann-Whitney U-test comparison of Summer 2026-vs-historical against historical-vs-historical median Bray-Curtis dissimilarity.

Supplemental Table 2: Mann-Whitney U test for rare-taxa per sample between host and non-host cities during the tournament window.

Supplemental Table 3: Mann-Whitney U test of Summer 2026 rare-taxa load to prior summers.

Supplemental Table 4: List of novel taxa sampled during the World Cup tournament sampling window.

## Notes

### Competing Interest Statement

The authors have declared no competing interest.

## References

Angelo KM. Mass gatherings. In: CDC Yellow Book: Health Information for International Travel. 2026 ed. Centers for Disease Control and Prevention; 2025. Accessed August 1, 2026. https://www.cdc.gov/yellow-book/hcp/travel-for-work-other/mass-gatherings.html

Bauer C, Reger N, Rustem H a L, et al. SeqBoard : a genomics-based data dashboard for comprehensive wastewater virome monitoring. Journal of the American Medical Informatics Association. 2026;33(8):1446–1456. doi:10.1093/jamia/ocag088

Chen S, Zhou Y, Chen Y, Gu J. fastp: an ultra-fast all-in-one FASTQ preprocessor. Bioinformatics. 2018;34(17):i884–i890. doi:10.1093/bioinformatics/bty560

Clark JR, Terwilliger A, Avadhanula V, Tisza M, Cormier J, Javornik-Cregeen S, Ross MC, Hoffman KL, Troisi C, Hanson B, Petrosino J, Balliew J, Piedra PA, Rios J, Deegan J, Bauer C, Wu F, Mena KD, Boerwinkle E and Maresso AW (2023) Wastewater pandemic preparedness: Toward an end-to-end pathogen monitoring program. Front. Public Health 11:1137881. doi: 10.3389/fpubh.2023.1137881

Clark JR, Maresso AW. Sewers to Solutions: A guide to wastewater pathogen monitoring. Annual Review of Medicine. 2025;77(1):493–508. doi:10.1146/annurev-med-062024-125121

Clark JR, Tisza MJ, Hammerquist AL, et al. Wastewater Parvovirus B19 Signal Amid Rising Maternal Cases. Preprint. medRxiv. 2025;2025.07.07.25331044. Published 2025 Jul 8. doi:10.1101/2025.07.07.25331044

Clark, J.R., Chirman, D., Prakash, H. et al. Statewide multi-year wastewater sequencing reveals dual origins of HIV-1 signal. Nat Commun 17, 7428 (2026). 10.1038/s41467-026-74140-7

Influenza (seasonal). World Health Organization. Accessed August 17, 2026. https://www.who.int/news-room/fact-sheets/detail/influenza-(seasonal).

Javornik Cregeen S, Tisza MJ, Hanson B, et al. Sequencing-Based Detection of Measles in Wastewater: Texas, January 2025. Am J Public Health. 2025;115(7):1115–1119. doi:10.2105/AJPH.2025.308146

Kendrick L. World Cup has economic impact on Texas [Internet]. Spectrum News 1 Texas; 2026 Jun 25 Available from: https://spectrumlocalnews.com/tx/south-texas-el-paso/news/2026/06/25/world-cup-has-economic-impact-on-texas

Lesenfants M, Suffredini E, Mancini P et al. Public health responses following identification of poliovirus in wastewater The Lancet Public Health, 2026; 11, e329–e342

Kakoullis L, Steffen R, Osterhaus A, et al. Influenza: seasonality and travel-related considerations. Journal of Travel Medicine. 2023;30(5). doi:10.1093/jtm/taad102

Manor Y, Handsher R, Halmut T, Neuman M, Bobrov A, Rudich H, Vonsover A, Shulman L, Kew O, Mendelson E 1999.Detection of Poliovirus Circulation by Environmental Surveillance in the Absence of Clinical Cases in Israel and the Palestinian Authority. J Clin

Kitajima M, Murakami M, Iwamoto R, Katayama H, Imoto S. Covid-19 wastewater surveillance implemented in the Tokyo 2020 olympic and Paralympic Village. Journal of Travel Medicine. 2022;29(3). doi:10.1093/jtm/taac004

Kuchinski, K. S., Loos, K. D., Suchan, D. M., Russell, J. N., Sies, A. N., Kumakamba, C., Muyembe, F., Kingebeni, P. M., Lukusa, I. N., N’Kawa, F., Losoma, J. A., Makuwa, M., Gillis, A., LeBreton, M., Ayukekbong, J. A., Lerminiaux, N. A., Monagin, C., Joly, D. O., Saylors, K., … Cameron, A. D. (2022). Targeted genomic sequencing with probe capture for discovery and surveillance of coronaviruses in bats. eLife, 11. 10.7554/elife.79777

Memish Z, Steffen R, White P et al. Mass gatherings medicine: public health issues arising from mass gathering religious and sporting events. The Lancet, 393, 2073–2084. doi: 10.1016/S0140-6736(19)30501-X

Prakash H, Perez RK, Ross M, Tisza M, Javornik Cregeen SJ, Deegan J, Petrosino JF, Boerwinkle E, Clark JR, Maresso AW. 2026. Detection, persistence, and rising prevalence of oncogenic viruses revealed by wastewater metagenomics. Appl Environ Microbiol 92:e00547–26.10.1128/aem.00547-26

Sullivan, N. J. (2026). Bundibugyo virus disease in 2026—clinical and public health responses. New England Journal of Medicine, 395(3), 278–289.

Sutton M, Radniecki TS, Kaya D, et al. Detection of SARS-CoV-2 B.1.351 (Beta) Variant through Wastewater Surveillance before Case Detection in a Community, Oregon, USA. Emerging Infectious Diseases. 2022;28(6):1101–1109. doi:10.3201/eid2806.211821.

Tisza, M., Javornik Cregeen, S., Avadhanula, V. et al. Wastewater sequencing reveals community and variant dynamics of the collective human virome. Nat Commun 14, 6878 (2023). 10.1038/s41467-023-42064-1

Tisza, M. J., Hanson, B. M., Clark, J. R., Wang, L., Payne, K., Ross, M. C., … & Maresso, A. W. (2024). Sequencing-based detection of avian influenza A (H5N1) virus in wastewater in ten cities. New England Journal of Medicine, 391(12), 1157–1159.

Toledo DM,Robbins AA, Gallagher TL, Hershberger KC, Barney RE, Salmela SM, Pilcher D, Cervinski MA, Nerenz RD, Szczepiorkowski ZM, Tsongalis GJ, Lefferts JA, Martin IW, Hubbard JA. 2022. Wastewater-Based SARS-CoV-2 Surveillance in Northern New England. Microbiol Spectr 10:e02207–21.10.1128/spectrum.02207-21

Ulhuq, F. R., Barge, M., Falconer, K., Wild, J., Fernandes, G., Gallagher, A., McGinley, S., Sugadol, A., Tariq, M., Maloney, D., Kenicer, J., Dewar, R., Templeton, K., & McHugh, M. P. (2023). Analysis of the ARTIC V4 and V4.1 SARS-CoV-2 primers and their impact on the detection of Omicron BA.1 and BA.2 lineage-defining mutations. Microbial genomics, 9(4), mgen000991. 10.1099/mgen.0.000991

